# The 2026 public charge rule and long-term health impacts among NYC immigrants: a simulation study

**DOI:** 10.64898/2026.07.23.26358686

**Authors:** Kaveri Nadhamuni, Evan Curcio, Sam Solomon, Sungwoo Lim, Gretchen Van Wye, Mamta Parakh

## Abstract

**Importance:** The 2026 public charge rule could discourage immigrants from accessing health coverage programs, creating a chilling effect that potentially leads to negative health outcomes; However, its long-term health impact is poorly understood.

**Objective:** To model potential impacts of the 2026 public charge rule on primary care and premature mortality among immigrants in New York City (NYC).

**Design, Setting, and Participants:** The simulation used a deterministic compartmental model with Ordinary Differential Equations (ODEs) using 2023 NYC Vital statistics data and American Community Survey, and estimates obtained from 2 previous studies about effects of healthcare access on primary care and Medicaid expansion on premature mortality.

**Main Outcomes and Measures:** Rates of primary care outcomes (access, doctor’s visits) in 5 years, and premature mortality in 5 and 20 years, projected by the model under conservative, moderate, and aggressive scenarios of avoidance/disenrollment due to the public charge rule, known as the “chilling effect”. Effects of the avoidance/disenrollment on primary care outcomes and premature mortality were obtained from 2 previous studies. Projected rates of the outcomes under each scenario were compared with counterfactuals to estimate the health impacts of the chilling effect.

**Results:** Implementation of the public charge rule was projected to decrease the primary care access rate by 4.1% (conservative) to 9.9% (aggressive) over 5 years, relative to the counterfactual scenario without the rule. The rate of doctors’ visits was projected to decrease over 5 years by 5.1% (conservative) to 12.2% (aggressive). Premature mortality was projected to increase by 4.4% (conservative) to 10.6% (aggressive) in 5 years and 7.4% (conservative) to 17.4% (aggressive) in 20 years. Legal noncitizens and Black immigrant New Yorkers were predicted to experience higher burdens of premature mortality attributed to the chilling effect, compared with other immigrant groups and racial/ethnic groups, respectively.

**Conclusions and Relevance:** This study demonstrates adverse health consequences of federal public charge rule changes among immigrants in NYC. The model projected a decrease in primary care visits and increase in premature mortality across various scenarios. These findings suggest urgent reconsideration of a regulatory change that disproportionately increases risk of premature mortality among immigrants in NYC.

**Key Points:** *Question:* What are the long-term impacts of the 2026 public charge rule on primary care and premature mortality among immigrants in New York City?

*Findings:* In this simulation study, under the scenario of a large-scale avoidance or disenrollment of public health services due to the rule vs. no such rule, the primary care access rate was projected to decrease by 10% over 5 years, and premature mortality rate was projected to increase by 17% in 20 years.

*Meaning:* The findings suggest urgent reconsideration of a regulatory change that has devastating public health consequences.

## Introduction

In July 2026, the U.S. Department of Homeland Security issued a new public charge inadmissibility rule, to repeal existing regulations and grant immigration officers greater discretionary authority to evaluate use of public benefits when reviewing immigration petitions. This rule will create uncertainty in immigration decisions, potentially discouraging immigrants from accessing public benefits, social services and essential health coverage programs such as Medicaid.^1^

Avoidance of public services due to fear of negative repercussions on their immigration petitions or on potential opportunity to seek immigration benefits, known as the “chilling effect,” was observed in 2019 after public charge regulations were modified.^2,3^ Even after the 2019 rule was briefly implemented and rescinded, 1 in 6 immigrant families with children reported avoiding noncash government benefits in the past year because of concerns about later obtaining a green card.^4^

Current evidence on the chilling effect is largely limited to observational evidence of healthcare utilization and coverage loss.^1^ We conducted a simulation study to model impacts of the public charge rule change on 2 leading health measures, primary care over 5 years and premature mortality over 20 years, among immigrant New York City (NYC) adults.

## Methods

We developed a deterministic compartmental model with Ordinary Differential Equations (ODEs) to predict the chilling effects on primary care visits and premature mortality. Our simulation population (N = 843,725) represented NYC immigrants aged 0 to 64 years, stratified into undocumented immigrants (UD), legal noncitizens (LNC), and children in mixed-status households (MSHC), using 2025 Center for Migration Studies data (Table A1, Appendix B).^5^ Compartments were defined by 6 dimensions: age group, immigration status, sex, race and ethnicity, insurance status, and chronic condition status.

The model governs 5 types of population flow. Insurance status transitions from Medicaid-enrolled to uninsured were modeled as an exponential decay concentrated in the first 3 years post-policy, calibrated to match each scenario’s cumulative loss. Baseline mortality used age- and race-specific rates from 2023 NYC Vital Statistics;^6^ excess mortality was applied to disenrolled individuals using an exposure-weighted hazard derived from published estimates of the premature mortality effect of loss of Medicaid.^7^ Aging was modeled as transitions between age strata at rates equal to the inverse of each age band’s width. Births were added to MSHC compartments, estimated from births to foreign-born mothers, scaled by the proportion of noncitizen foreign-born New Yorkers.^6^ Net immigrant flows incorporated domestic and international migration of noncitizens from American Community Survey (ACS) Public Use Microdata and naturalization estimates over the prior decade.^5^ Together, these yield the system of ODEs (Figure 1, Appendix C).

**Figure 1.**
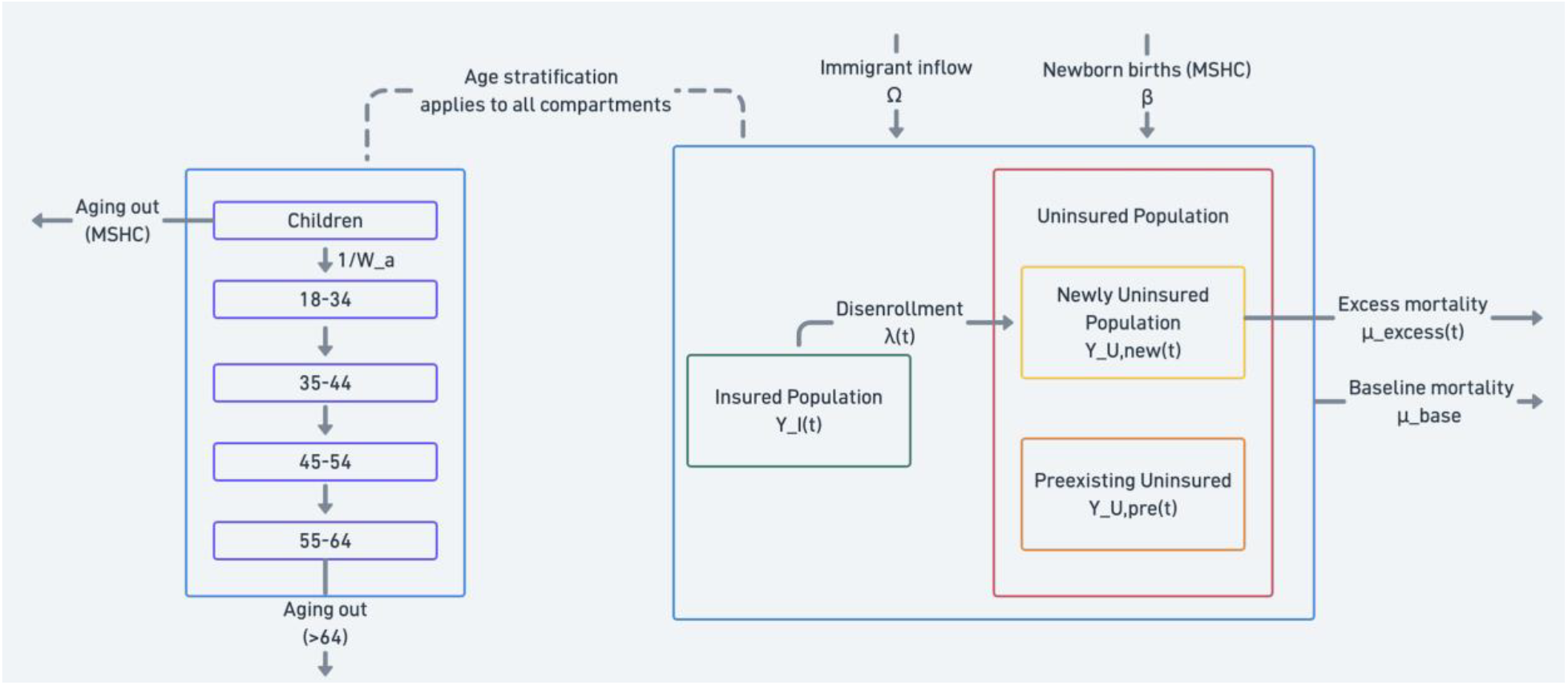
Schematic of the compartmental model and state transitions Notes: The high-level compartmental overview (**right**) illustrates the population structure and dynamics. Additive inputs to the system include newborn births, β, flowing into MSHC (Mixed-Status Household Children) and immigrant inflow, Ω. It isolates the policy-driven transition from the Insured Population, Y_I_(t), to the Newly Uninsured Population, Y_U,new_(t), via the disenrollment rate, λ(t). Baseline mortality, μ_base_(t), applies to the general population, whereas excess mortality, μ_excess_(t), is restricted strictly to the newly uninsured cohort. Population flows across compartments is driven by these insurance transitions and aging flows (**left**). Within this aging mechanism, individuals transition sequentially between age bands at rates proportional to the inverse of the band’s width, 1/W_a_. Exits from the simulation via aging occur at two specific boundaries: children within the MSHC compartments age out of the simulation upon reaching adulthood, and adults in the oldest age band age out upon exceeding age 64.

Using the 2025 KFF/New York Times Survey of Immigrants data,^8^ we modeled 4 scenarios for disenrollment:

1. Baseline: no avoidance/disenrollment, as a control
2. Conservative: 10% avoidance/disenrollment for LNC and 25% for UD
3. Moderate: 20% avoidance/disenrollment for LNC and 35% for UD
4. Aggressive: 30% avoidance/disenrollment for LNC and 42% for UD.

Primary care parameters came from a randomized controlled study among uninsured immigrant New Yorkers.^9^ Uninsured and insured compartments were assigned control-group and post-enrollment rates, respectively. Excess premature mortality from avoidance/disenrollment was derived from quasi-experimental literature leveraging the Affordable Care Act’s Medicaid expansion as a natural experiment.^7, 10-12^ We scaled state-level data using a duration-dependent saturation function fit separately to adults aged 55–64 years and adults aged 19–54 years, with plateau constraints informed by evidence across multiple studies.^7, 10-12^

Projected rates of outcomes under each scenario were then compared with counterfactuals (i.e., rates under baseline scenario) in 5- and 20-years post-policy, and rate differences were reported as chilling effects on outcomes.

Model code is available at https://github.com/nychealth/public-charge-simulation.

## Results

In Year 0, 84% of LNC adults and 42% of UD adults were enrolled in public insurance (Figure D1). Over 5 years, LNC adults were predicted to avoid healthcare or drop coverage due to the chilling effect by a rate of 10% (conservative) to 30% (aggressive). A higher rate of decrease was predicted among UD adults (conservative: 25%; aggressive: 42%).

The public charge rule was projected to reduce the PCP access rate by 4.1% (conservative) to 9.9% (aggressive) over 5 years compared with the baseline scenario (Table 1). Similarly, the rate of doctors’ visits was projected to decrease over 5 years by 5.1% (conservative) to 12.2% (aggressive). Reductions in PCP access and doctors’ visits attributed to the chilling effect did not differ substantially across immigrant subgroups (Tables E1).

**Table 1.** Projected rates of primary care provider access and doctors’ visits per 1000 immigrant New Yorkers, during 5 years after the public charge rule. Notes: baseline = no avoidance/disenrollment, as a control; conservative = 10% avoidance/disenrollment for legal noncitizens and 25% for undocumented; moderate = 20% avoidance/disenrollment for legal noncitizens and 35% for undocumented; aggressive = 30% avoidance/disenrollment for legal noncitizens and 42% for undocumented

| Scenario | Year 0 rate | Projected Year 5 rate | Difference from Baseline in Year 5 rate | % change from Baseline in Year 5 rate |
| --- | --- | --- | --- | --- |
| Primary care provider access per 1000 |  |  |  |  |
| Baseline | 384.9 | 385.8 | - | - |
| Conservative | 384.9 | 370.0 | -15.9 | -4.1% |
| Moderate | 384.9 | 358.5 | -27.4 | -7.1% |
| Aggressive | 384.9 | 347.8 | -38.1 | -9.9% |
| Doctors' visits per 1000 |  |  |  |  |
| Baseline | 1616.3 | 1620.1 | - | - |
| Conservative | 1616.3 | 1537.9 | -82.3 | -5.1% |
| Moderate | 1616.3 | 1478.5 | -141.7 | -8.7% |
| Aggressive | 1616.3 | 1422.8 | -197.3 | -12.2% |

Baseline = no avoidance/disenrollment, as a control; conservative = 10% avoidance/disenrollment for legal noncitizens and 25% for undocumented; moderate = 20% avoidance/disenrollment for legal noncitizens and 35% for undocumented; aggressive = 30% avoidance/disenrollment for legal noncitizens and 42% for undocumented.

Premature mortality was projected to increase by 4.4% (conservative) to 10.6% (aggressive) in 5 years and 7.4% (conservative) to 17.4% (aggressive) in 20 years, compared with no public charge rule scenario. Of racial and ethnic subgroups, Black immigrant New Yorkers were predicted to experience a slightly larger increase in premature mortality than other groups in both 5 and 20 years (Figure 2, Table F3). Under the conservative scenario, the percent changes in premature mortality, relative to no such rule, were slightly lower among LNC adults than UD adults (5 years: 3.6% vs. 5.1%, 20 years: 6.7% vs. 8.0%). However, higher chilling effects on premature mortality were predicted among LNC versus UD adults under moderate (5 years: 7.7% vs. 7.6%, 20 years: 13.9% vs. 11.0%) and aggressive scenarios (5 years: 12.4% vs. 9.1%, 20 years: 20.6% vs. 13.0%) (Figure 2, Table F2).

**Figure 2.**
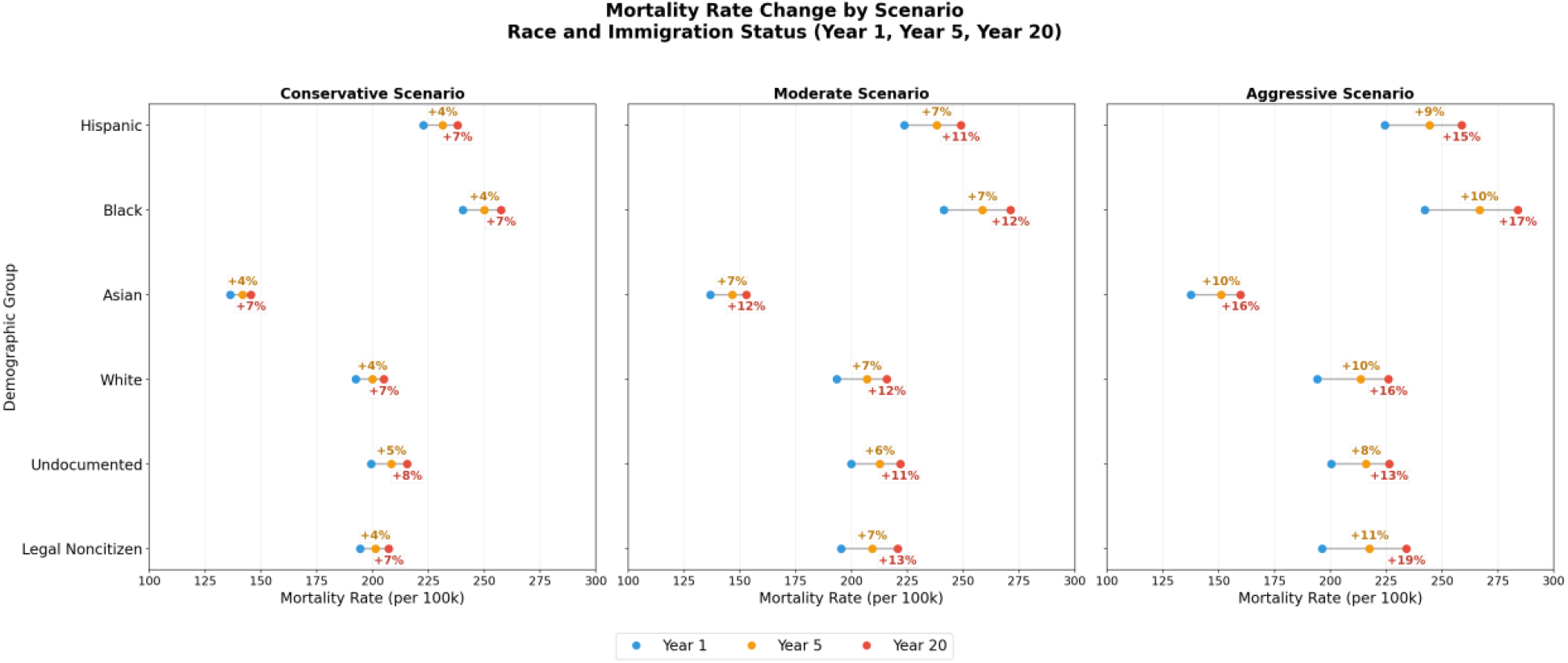
Change in Premature Mortality Rate in various scenarios, by race/ethnicity and immigrant status. Notes: Points correspond to premature mortality rate at respective years (Year 1, blue marker; Year 5, yellow marker; Year 20, red marker), and percentages correspond to relative change under each scenario compared with baseline scenario in Year 5 (yellow, above) and Year 20 (red, below), respectively.

## Discussion

In this simulation study, we found that the 2026 public charge rule could substantially reduce primary care utilization and increase premature mortality among immigrant New Yorkers. Under the scenario of large-scale avoidance/disenrollment due to the public charge rule versus no such rule, PCP access and doctors’ visit rates were projected to decline by 9.9% and 12.2% within 5 years, respectively. Under the same scenario, premature mortality rates were projected to increase by 10.9% in 5 years, 17.4% in 20 years, relative to no such rule, consistent with avoidance/disenrollment effects in Medicaid expansion literature.^7^ Further, due to the chilling effect, legal noncitizens and Black immigrant New Yorkers were predicted to experience higher burdens of premature mortality within their respective populations. These findings are consistent with and extend prior literature by modeling premature mortality impacts over a 20-year horizon.^2,3^

## Limitations

We could not quantify international outflow because the ACS only surveys U.S. residents, potentially leading to overestimation of immigrant inflow. The premature mortality effects were derived from the Miller et al. study^7^ of U.S. citizens across many states, and we assumed coverage loss produces symmetric opposite effects (i.e., same effects in the opposite direction), which is theoretically plausible but untested. The effects of the public charge rule are difficult to separate from broader immigration policies (e.g., immigration enforcement). Our simulation draws on studies of the 2019 public charge enforcement and the 2025 benefits avoidance surveys, which show higher avoidance from healthcare.^2.3^ Our results likely reflect the combined impact of multiple policies and heightened immigration-related concern rather than any single policy alone. To account for uncertainty in policy implementation, we employed 3 scenarios spanning conservative to aggressive assumptions, yielding a range of estimates.

## Conclusions

This study demonstrates adverse health consequences of the federal public charge rule changes among immigrant New Yorkers. The model projected a decrease in primary care visits and increase in premature mortality across various scenarios. These findings suggest an urgent need for reconsideration of a regulatory change that will disproportionately increase the risk of dying early among immigrant New Yorkers.

## Data Availability

The simulation code and associated data are available through the public GitHub repository provided in the Methods section of the manuscript. The repository contains the complete implementation of the simulation, associated data files, and output tables and figures. Parameter tables and demographic tables are also provided in the supplementary materials.

https://github.com/nychealth/public-charge-simulation

## Acknowledgements

Claude Sonnet version 4.6 via GitHub Copilot (Anthropic, Microsoft) was used to support background research, and coding assistance for the simulation model. All AI-generated suggestions were reviewed, validated, and modified by the authors, who take responsibility for the integrity and accuracy of the final code.

## References

1. Bustamante AV, Barajas CB, Ortega AN. The Public Health Consequences of the 2025 Public Charge Announcements—Uncertainty as Policy. JAMA Network Open. 2026 Jan 29;9(1):e2555044.

2. Alulema, Daniela and Jacquelyn Pavilon. 2022. Immigrants’ Use of New York City Programs, Services, and Benefits: Examining the Impact of Fear and Other Barriers to Access. Center for Migration Studies of New York (CMS) Report. New York, NY: CMS.

3. Ettinger de Cuba S, Miller DP, Raifman J, Cutts DB, Bovell-Ammon A, Frank DA, Jones DK. Reduced health care utilization among young children of immigrants after Donald Trump’s election and proposed public charge rule. Health Affairs Scholar. 2023 Aug;1(2):qxad023.

4. Gonzalez, D., Haley, J. M., & Kenney, G. (2023, November). One in six adults in immigrant families with children avoided public programs in 2022 because of green card concerns. Urban Institute. https://www.urban.org/research/publication/one-six-adults-immigrant-families-children-avoided-public-programs-2022.

5. Center for Migration Studies. Data briefing: a portrait of immigrant New Yorkers. Published November 14, 2025. Accessed December 1, 2025. https://cmsny.org/publications/data-briefing-on-new-york-city-immigrants/.

6. Li W, Seil K, Paudel P, et al. Summary of Vital Statistics, 2023. New York, NY: Bureau of Vital Statistics, New York City Department of Health and Mental Hygiene; 2024.

7. Miller S, Johnson N, Wherry LR. Medicaid and mortality: new evidence from linked survey and administrative data. The Quarterly Journal of Economics. 2021 Aug;136(3):1783–829.

8. Pillai D, Artiga S, Pillai A, et al. KFF/New York Times 2025 Survey of Immigrants: health and health care experiences during the second Trump administration. KFF. Published November 18, 2025. Updated February 11, 2026. Accessed February 17, 2026. https://www.kff.org/immigrant-health/kff-new-york-times-2025-survey-of-immigrants-health-and-health-care-experiences-during-the-second-trump-administration/

9. Gruber J, Sabety A, Sood R, Bae JY. Reducing frictions in healthcare access: the ActionHealth NYC experiment for undocumented immigrants. National Bureau of Economic Research; 2022 Mar 14.

10. Wilper AP, Woolhandler S, Lasser KE, McCormick D, Bor DH, Himmelstein DU. Health insurance and mortality in US adults. American journal of public health. 2009 Dec;99(12):2289–95.

11. Sommers BD, Baicker K, Epstein AM. Mortality and access to care among adults after state Medicaid expansions. New England Journal of Medicine. 2012 Sep 13;367(11):1025–34.

12. Wyse A, Meyer BD. Saved by Medicaid: new evidence on health insurance and mortality from the universe of low-income adults. National Bureau of Economic Research; 2025 May 5.

