## Appendix for "The 2026 public charge rule and long-term health impacts among NYC immigrants: a simulation study"

### Appendix A. Demographic characteristics of simulation population

Table A1. Demographic characteristics of the synthetic population

| | Undocumented (UD) | Legal non-citizen (LNC) |
|---|---|---|
| **Sex** | | |
| Female | 46% | 54% |
| Male | 54% | 46% |
| **Race/ethnicity** | | |
| Hispanic | 50% | 37% |
| Asian non-Hispanic | 24% | 28% |
| Black non-Hispanic | 11% | 13% |
| White non-Hispanic | 10% | 19% |
| Other non-Hispanic | 5% | 4% |
| **Medicaid coverage**[a] | | |
| Adults | 25% | 47% |
| Children | 64% | 61% |
| **Uninsured**[b] | | |
| Adults | 34% | 9% |
| Children | 9% | 4% |
| **Age groups** | | |
| <18 years | 18% | 9% |
| 18-34 years | 33% | 37% |
| 35-44 years | 18% | 20% |
| 45-54 years | 16% | 17% |
| 55-64 years | 15% | 17% |

Notes:

[a]% did not add up to 100% since only % of those with Medicaid were reported.

[b]% did not add up to 100% since only % of those without insurance were reported.

**Appendix B.**

This report's use of the term "undocumented" is broadly defined, including categories of individuals with various forms of temporary or interim immigration status, pending applications, and similar status, sometimes referred to as "permanently residing under color of law" or liminal statuses. A substantial proportion of this population is eligible for public health insurance programs pursuant to New York State courts' construction of the state's Constitution or, for those with employment authorization based on their status, for employment-sponsored insurance. In addition, some "undocumented" immigrants may have insurance coverage as dependents of an insured family member.

**Appendix C. Model Equations and Parameters**

1. Insurance transitions from Medicaid enrolled to uninsured are governed by:
$\frac{dY_I}{dt} = -\lambda(t) \cdot Y_I(t), \frac{dY_{U,\,new}}{dt} = \lambda(t) \cdot Y_I(t),$
where $Y_I$ and $Y_{U,\,new}$ are the insured and newly uninsured populations, respectively. The scenario-specific disenrollment hazard is defined as $\lambda(t) = \left(A \cdot e^{-t/T_{decay}}\right)$, where:
$$A = \frac{-\ln\left(1-\phi_{i,s}\right)}{T_{decay}\left(1-e^{-T_{horizon}/T_{decay}}\right)}$$
2. Baseline mortality is applied to the insured and pre-existing uninsured populations:
$\frac{dY_I}{dt} = -\mu_{base}(t) \cdot Y_I(t)\,, \frac{dY_{U,\,pre}}{dt} = -\mu_{base}(t) \cdot Y_{U,\,pre}(t)$ where $Y_{U,\,pre}$ is the preexisting uninsured population from the beginning of the simulation. Excess mortality is added additionally to the newly uninsured population:
$\frac{dY_{U,\,new}}{dt} = -\left(\mu_{base}(t) + \mu_{excess}(t)\right) \cdot Y_{U,\,new}(t),$
Excess mortality $\mu_{excess}(t) = \frac{f\left(\tau(t)\right)}{\mu_{ref} \cdot \Delta_{ref}} \cdot \mu_{base}(t)$, where the mortality saturation of the form $f(t) = a \cdot (1 - e^{-bt}) + c$ is fitted to Miller et al.'s study.[7]
3. Continuous aging across sequential age bins:
$\frac{dY_{a,imm}}{dt} = \mathrm{I}(a > 1) \cdot \mathrm{I}(imm \neq MSHC) \cdot \frac{1}{W_{a-1}} \cdot Y_{a-1,imm}(t) - \frac{1}{W_a} \cdot Y_{a,imm}(t),$
for any compartment with age index $a \in \{1,2,3,4,5\}$ and immigration status, *imm*.
4. Births added to MSHC compartments: $\frac{dY_{children,MSHC}}{dt} = \beta \cdot \Phi_{r,g,n,h}$ The outflow term removes MSHC children from the simulation upon reaching age 18.
5. Annual immigrant inflow is distributed across non-MSHC compartments, matching the current compartment distribution $\frac{dY_{imm}}{dt} = \Omega \cdot \omega_{imm}\left(Y(t)\right)\ \forall\ imm \notin MSHC$

Table C1. ODE Parameters

| ODE parameters | Estimates | Notes |
|---|---|---|
| $\phi_{i,s}$ | | Scenario and immigration status-specific disenrollment fraction |
| $T_{decay}$ | 3.0 | The exponential decay constant for the disenrollment hazard function. |
| $T_{horizon}$ | 100 | The integration cutoff used to ensure that the total disenrollment fraction integrates correctly to its target value. |
| $\mu_{base}(t)$ | | Baseline annual mortality rate stratified by age group and race/ethnicity, derived from foreign born rates from NYC Vital Statistics 2023 Report |
| $\mu_{excess}(t)$ | | Excess mortality due to policy disenrollment, applied exclusively to newly disenrolled individuals. It is modeled as a time-dependent hazard driven by an exponential saturation function of exposure time. |
| $\tau(t)$ | | Average number of years spent uninsured for the newly uninsured in a compartment at time t. |
| $\mu_{ref}$ | Ages 18-54: 430 (per 100,000)<br>Ages 55-64: 1,400 (per 100,000) | The reference baseline mortality rate for an age group, used to calibrate the Miller et al. effect. |
| $\Delta_{ref}$ | Ages 18-54: 13.4%<br>Ages 55-64: 14.7% | The Medicaid coverage change for an age group, from Miller et al. |
| $W_a$ | Ages 0-17 (Children): 18<br>Ages 13-34: 17<br>Ages 35-44: 10<br>Ages 45-54: 10<br>Ages 55-64: 10 | The width of each age group bin, used to compute aging transition rates |
| $\Phi_{r,g,n,h}$ | | Proportional distribution factor across race/ethnicity (r), gender (g), insurance status (n), and health status (h), derived from demographic |

| | | |
|---|---|---|
| | | data to allocate births across MSHC compartments. |
| β | 19,672 | Total annual births to foreign-born noncitizen mothers derived from NYC Vital Statistics 2023 Report |
| Ω | 18,000 | Annual immigrant inflow to NYC estimated from ACS PUMS domestic and international migration and naturalization rates |
| $\omega_{imm}$ | | Distribution factor allocating annual immigrant inflow Ω across non-MSHC compartments, matched to current compartment distribution at each time step. |

**Appendix D.**

Figure D1. Projected annual trend of insurance enrollment during 5 years after the public charge rule among immigrant New Yorkers

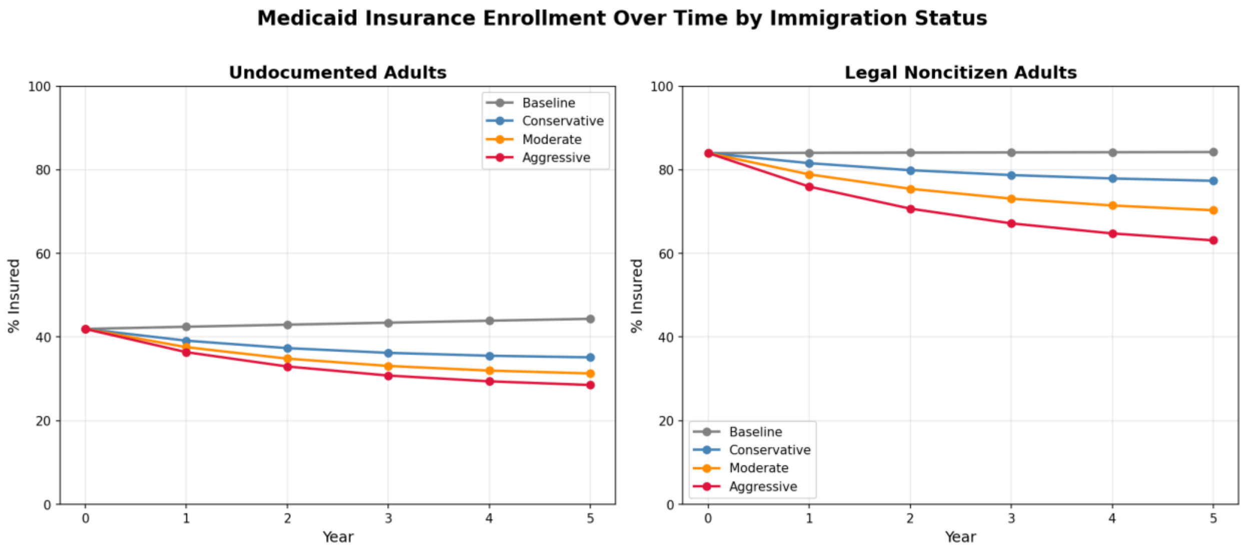

Notes: baseline = no avoidance/disenrollment, as a control; conservative = 10% avoidance/disenrollment for legal noncitizens and 25% for undocumented; moderate = 20% avoidance/disenrollment for legal noncitizens and 35% for undocumented; aggressive = 30% avoidance/disenrollment for legal noncitizens and 42% for undocumented.

**Appendix E.**

Table E1. Rates of doctors' visits and primary care physician (PCP) access per 1000 among Immigrants in New York City from year 0 to year 5 by immigration status

| **Scenario** | **Year 0 rate** | **Year 5 rate** | **Difference from baseline in Year 5 rate** | **% change from baseline in Year 5 rate** |
|---|---|---|---|---|
| PCP access per 1000 | | | | |
| Undocumented | | | | |
| Baseline | 335.2 | 339.0 | | -- |
| Conservative | 335.2 | 320.5 | -18.5 | -5.5% |
| Moderate | 335.2 | 312.8 | -26.2 | -7.7% |
| Aggressive | 335.2 | 307.2 | -31.8 | -9.4% |
| Legal non-citizen | | | | |
| Baseline | 420.7 | 420.6 | | -- |
| Conservative | 420.7 | 406.7 | -13.9 | -3.3% |
| Moderate | 420.7 | 392.4 | -28.2 | -6.7% |
| Aggressive | 420.7 | 377.8 | -42.8 | -10.2% |
| Doctors' visits per 1000 | | | | |
| Undocumented | | | | |
| Baseline | 1358.6 | 1376.6 | | -- |
| Conservative | 1358.6 | 1280.9 | -95.7 | -7.0% |
| Moderate | 1358.6 | 1241.0 | -135.6 | -9.9% |
| Aggressive | 1358.6 | 1212.4 | -164.3 | -11.9% |
| Legal non-citizen | | | | |
| Baseline | 1802.1 | 1801.0 | | -- |
| Conservative | 1802.1 | 1728.7 | -72.3 | -4.0% |
| Moderate | 1802.1 | 1654.8 | -146.1 | -8.1% |
| Aggressive | 1802.1 | 1579.2 | -221.8 | -12.3% |

Notes (apply to tables E1 to F3): baseline = no avoidance/disenrollment, as a control; conservative = 10% avoidance/disenrollment for legal noncitizens and 25% for undocumented; moderate = 20% avoidance/disenrollment for legal noncitizens and 35% for undocumented; aggressive = 30% avoidance/disenrollment for legal noncitizens and 42% for undocumented.

**Appendix F. Mortality Outcomes**

Table F1. Overall Premature Mortality Rates among Immigrants in New York City

| Scenario | Year 1 | Year 5 | Year 20 |
|---|---|---|---|
| Baseline | 19.6 (REF) | 19.6 (REF) | 19.7 (REF) |
| Conservative | 19.6 (+0.5%) | 20.4 (+4.5%) | 21.1 (+7.4%) |
| Moderate | 19.7 (+0.9%) | 21.1 (+7.7%) | 22.1 (+12.6%) |
| Aggressive | 19.8 (+1.3%) | 21.7 (+10.9%) | 23.1 (+17.4%) |

Table F2. Premature Mortality Rates among Immigrants in New York City by Immigration Status

| Scenario | Immigration Status | Year 1 | Year 5 | Year 20 |
|---|---|---|---|---|
| Baseline | Undocumented | 19.8 (REF) | 19.8 (REF) | 20.0 (REF) |
| Conservative | | 19.9 (+0.5%) | 20.8 (+5.1%) | 21.6 (+8.0%) |
| Moderate | | 20.0 (+1.0%) | 21.3 (+7.6%) | 22.2 (+11.0%) |
| Aggressive | | 20.0 (+1.0%) | 21.6 (+9.1%) | 22.6 (+13.0%) |
| Baseline | Legal non-Citizen | 19.4 (REF) | 19.4 (REF) | 19.4 (REF) |
| Conservative | | 19.4 (+0.0%) | 20.1 (+3.6%) | 20.7 (+6.7%) |
| Moderate | | 19.5 (+0.5%) | 20.9 (+7.7%) | 22.1 (+13.9%) |
| Aggressive | | 19.6 (+1.0%) | 21.8 (+12.4%) | 23.4 (+20.6%) |

Table F3. Premature Mortality Rates among Immigrants in New York City by Race and Ethnicity

| Scenario | Race/Ethnicity | Year 1 | Year 5 | Year 20 |
|---|---|---|---|---|
| Baseline | Hispanic | 22.2 (REF) | 22.2 (REF) | 22.2 (REF) |
| Conservative | | 22.3 (+0.5%) | 23.1 (+4.1%) | 23.8 (+7.2%) |
| Moderate | | 22.4 (+0.9%) | 23.8 (+7.2%) | 24.9 (+12.2%) |
| Aggressive | | 22.4 (+0.9%) | 24.4 (+9.9%) | 25.9 (+16.7%) |
| Baseline | Black | 23.9 (REF) | 23.9 (REF) | 23.9 (REF) |
| Conservative | | 24.0 (+0.4%) | 25.0 (+4.6%) | 25.8 (+7.9%) |
| Moderate | | 24.1 (+0.8%) | 25.9 (+8.4%) | 27.1 (+13.4%) |
| Aggressive | | 24.2 (+1.3%) | 26.7 (+11.7%) | 28.4 (+18.8%) |
| Baseline | Asian | 13.6 (REF) | 13.6 (REF) | 13.6 (REF) |
| Conservative | | 13.6 (+0.0%) | 14.2 (+4.4%) | 14.6 (+7.4%) |
| Moderate | | 13.7 (+0.7%) | 14.7 (+8.1%) | 15.3 (+12.5%) |
| Aggressive | | 13.7 (+0.7%) | 15.1 (+11.0%) | 16.0 (+17.6%) |
| Baseline | White | 19.2 (REF) | 19.2 (REF) | 19.2 (REF) |
| Conservative | | 19.3 (+0.5%) | 20.0 (+4.2%) | 20.5 (+6.8%) |
| Moderate | | 19.3 (+0.5%) | 20.7 (+7.8%) | 21.6 (+12.5%) |
| Aggressive | | 19.4 (+1.0%) | 21.4 (+11.5%) | 22.6 (+17.7%) |
